# The overlap between frailty and dementia, and impact on hospitalisation and mortality

**DOI:** 10.64898/2026.11.17.26368281

**Authors:** Yolanda Lau, Claudia Cooper, Ruth Dobson, Sube Banerjee, Harriet Demnitz-King, DeNPRU-QM

## Abstract

**Background:** Dementia and frailty are common age-related syndromes that frequently coexist and independently increase the risk of hospitalisation and mortality. Whether frailty confers additional risk among people with dementia, compared with those without dementia, is unclear. We examined the prevalence of frailty among people with dementia and its association with hospitalisation and mortality in England.

**Methods:** Using English primary care data from the Clinical Practice Research Datalink, linked to hospital and mortality records, we matched people with dementia to up to ten controls without dementia by year of birth, sex, and practice. Frailty was measured using the electronic Frailty Index. Outcomes were all-cause hospitalisation (episodes and total bed days) and mortality, analysed using negative binomial, and Cox proportional hazards models. Interaction terms examined whether associations were modified by ethnicity, deprivation, and rurality.

**Results:** Among 335,172 people with dementia and 3,065,858 matched controls alive in 2021, 58.1% of people with dementia were living with frailty (30.5% mild, 19.2% moderate, 8.4% severe), compared with 29.4% of matched controls (21.7% mild, 6.3% moderate, 1.5% severe). To examine the impact of frailty on outcomes, we used a separate analytical sample of 556,685 people with dementia and 1,526,612 matched controls, followed from index date (dementia diagnosis for cases; matched controls assigned the same dates). Follow-up period was 4.1 years in people with dementia and 6.8 years in matched controls. In analyses that controlled for follow-up time, among people with dementia, relative to the control group without frailty, greater frailty severity was associated with more hospitalisation episodes (severe frailty aIRR = 1.47, 95% CI 1.45–1.50, p<0.001) and higher mortality (aHR = 1.58, 95% CI 1.54–1.62, p<0.001), while mild and moderate frailty were associated with fewer (aIRR = 0.96, 95% CI 0.95–0.97, p<0.001; 0.99 (0.97, 0.99) p=0.002) and severe frailty a greater number of bed days (aIRR = 1.04, 95% CI 1.01-1.10, p<0.001). Associations were similar but stronger among controls (severe frailty: episodes aIRR= 3.07, 95% CI 2.99–3.14; total bed days aIRR = 3.28, 95% CI 3.17–3.38; mortality aHR = 2.74, 95% CI 2.66–2.82; all p<0.001).

**Conclusions:** Frailty is highly prevalent among people with dementia and associated with more hospital admissions and mortality. The relationship with number of bed days is more complex, with longer duration in those with dementia without frailty than mild or moderate frailty, perhaps reflecting the complexity of discharge to independent living versus residential care settings. The assessment and management of frailty should be considered as a core part of dementia care.

## INTRODUCTION

Global populations are ageing^1^, and a growing number of older adults are living with age-related conditions, including dementia^2^ and frailty^3^. Frailty is a long-term condition characterised by vulnerability to adverse health outcomes; it exists on a spectrum of severity and often presents non-specifically through functional decline. International evidence reports a wide range of prevalence estimates, reflecting differences in populations, definitions and measurement approaches^4^.

Frailty and dementia frequently coexist and may interact to increase the risk of adverse outcomes. Frailty is associated with an increased risk of dementia^5^ and is common among people living with dementia^6^. In England, approximately 676,000 people have dementia^7^, and more than 1.5 million people aged 65 and over are living with frailty^8^. A systematic review found that over half of people with dementia in acute care settings, and 25% to 99% of those in the community, met criteria for frailty^9^.

A 2026 systematic review reported that frailty in people with dementia was associated with higher mortality, hospitalisation, and institutionalisation, and poorer physical function and quality of life^6^. However, the review included only 12 studies, none conducted in the UK, and few compared outcomes against a matched controls without dementia^6^. Among people with dementia, mortality^10^ and healthcare use^11^ have been found to vary by health inequality, including deprivation, ethnicity, and rurality. Similarly, greater frailty has been associated with higher deprivation, certain ethnic groups, and urban dwelling^12^. Whether the relationship between frailty and outcomes in people with dementia varies by ethnicity, deprivation, or rurality has received little attention.

In the 2025 NHS Ten Year Health Plan, the English government committed to developing modern service frameworks to set national standards and reduce unwarranted variation in care^13^. The development of the Modern Service Framework which covers both frailty and dementia is underway, yet how the two conditions co-occur, and how frailty transfigures health outcomes for people with dementia, remains largely unexamined.

Using population-based primary care data from England, we aimed to address three questions: (1) what proportion of people with a dementia diagnosis in England were living with different levels of frailty in 2021; (2) how all-cause hospitalisation and mortality rates differ between people with dementia and matched controls, according to baseline frailty severity; and (3) whether these relationships differ by ethnicity, deprivation, and rurality.

## METHODS

### Study design

This was a population-based matched study using primary care data from England, including people with dementia and matched counterparts without dementia.

### Data sources

Our data source was general practices in England contributing to the Clinical Practice Research Datalink (CPRD) GOLD and AURUM databases between 1 January 2000 and 31 December 2023. CPRD contains anonymised data on approximately 60 million patients from GP practices across the UK, including around 18 million who are currently registered^14^. We linked these data to Hospital Episode Statistics (HES) datasets for National Health Service (NHS) secondary care in England, and Office for National Statistics (ONS) death registration. For area-level data, we utilised the 2011 Rural-Urban Classification,based on the patient’s registered practice, and the Index of Multiple Deprivation (IMD), an area-level measure calculated at Lower Layer Super Output Area (LSOA) level. Data extraction was conducted separately for GOLD and AURUM. Data from GOLD and AURUM were then combined, with deduplication of GOLD records.

### Study population

We included patients who were registered to a primary care practice in England between between 1 January 2000 and 31 December 2023, with a recorded dementia diagnosis (using verified clinical code lists^15,16^ and in consultation with clinicians) who were eligible for linkage across all datasets, and their matched controls.

For all analyses, we excluded patients with no recorded year of birth, practice registration date, or dementia diagnosis date; those aged outside 50–105 years at diagnosis; and dementia cases without a confirmed or suspected diagnostic or prescription code, removed together with their matched controls.

Our analytic sample for research question 1 comprised all individuals alive with a recorded dementia diagnosis on or before 31 December 2021, and those without a dementia diagnosis, contributing data in 2021. We excluded individuals not registered with their general practice for at least 12 months during the study period, to allow sufficient time for data to be recorded.

Research questions 2 and 3 analytic samples comprised people with dementia followed from their index date (of diagnosis) and matched controls. We excluded those with a first dementia code recorded within six months of registration with their GP practice, as this may reflect information transfer rather than a true first diagnosis; those with less than two years of follow-up after diagnosis; and with missing covariate data. To identify outcomes, a six-month wash-out period was applied before and after the diagnosis, to ensure that outcomes did not arise from the process of diagnosis itself.

#### Dementia cases

Dementia was defined by the presence of a diagnostic code (Supplementary Table S1) or prescription code for dementia treatments (Supplementary Table S2). This list is more specific than the extensive code list used for data extraction. The date of the first recorded code was taken as the date of initial diagnosis, at which point people with dementia entered the study (index date).

#### Matched controls

As part of the dataset generation, each person with dementia was randomly matched to up to 10 controls without dementia by sex, year of birth and practice. CPRD do not permit access to the entire dataset, therefore this matching was performed before exclusion criteria were applied. Controls were assigned the index date of their matched person with dementia.

### Exposure

Baseline frailty was measured using the validated electronic Frailty Index (eFI)^17^, a deficit accumulation measure based on the presence or absence of 36 health deficits (35 deficits plus polypharmacy, defined as five or more medications in the previous 12 months) recorded in primary care (Supplementary Table S3). The score is the proportion of deficits present (number of deficits divided by 36), with higher scores indicative of greater frailty.

Baseline frailty was calculated using all available records up to the baseline date (the index date for the outcome analyses, and 31 December 2021 for RQ1). To ascertain deficits up to that point, and in line with published guidance, no lookback restriction was applied to individual deficits – each deficit was counted as present if ever recorded on or before the assessment date – with the exception of polypharmacy, which was defined over the preceding 12 months.

Frailty was categorised into four severity levels using established cut-points: no frailty (0–0.12), mild (>0.12–0.24), moderate (>0.24–0.36), and severe (>0.36) frailty^17^.

### Outcomes

We ascertained hospitalisations through linkage to HES APC. For each participant over the follow-up period, we derived the number of admissions (hospitalisation episodes) for any cause and the total number of days spent in hospital (total bed days). For all-cause mortality, we extracted dates of death through linkage to ONS death registration.

### Covariates

Covariates included age group (5-year bands: <60, 60–64, 65–69, 70–74, 75–79, ≥80), sex, ethnicity (self-reported using UK Census categories), smoking status, urban/rural classification (based on the registered practice), and IMD (an area-level measure of relative deprivation calculated at the Lower-layer Super Output Areas level)^18^. We did not adjust for individual comorbidities, as some these are components of the eFI.

### Missing data

Those with missing age, sex, IMD, or rural/urban classification were excluded; missing ethnicity data were retained as “unknown”; and missing smoking status was imputed. Differences (t-test or Mann-Whitney U test) between those with and without missing data were examined to assess whether missingness was plausibly consistent with a missing-at-random assumption. Significant differences were observed between those with and without smoking status.

Smoking status was imputed using multiple imputation by chained equations, using five imputed datasets.

### Statistical analysis

For research question one, we stratified findings by age group and sex, using chi-squared to test whether frailty levels differ between individuals with dementia and without dementia.

For research question two, baseline was defined as the index date – of dementia diagnosis for people with dementia, and the same assigned date for their matched controls – and participants were followed until death or the end of the study period. We fitted unadjusted and adjusted models using negative binomial regressions for hospitalisation episodes and total bed days, and Cox proportional hazards regressions for mortality. For total bed days, a log person-years offset was included so that estimates represent bed days per person-year, accounting for differences in follow-up duration between participants and the two cohorts. We assessed the proportional hazards assumption for the Cox models using Schoenfeld residuals. Adjusted models included ethnicity, IMD, rural/urbanity, smoking status, age, and sex, with random effects to account for variation between practices. Results were presented as hazard ratios with 95% confidence intervals. We included an interaction term between dementia status and frailty category. Outcomes were compared across frailty groups (mild, moderate, and severe frailty). As matching was performed before exclusions, the final sample carried small residual imbalances in age and sex. To examine whether these affected our estimates, in sensitivity analyses, we repeated analyses in the subset of matched sets that retained at least one control.

For research question three, we added interaction terms to models to examine whether inequalities (ethnicity, IMD, and rurality) modified the examined associations, using a likelihood ratio test. Results were stratified by the interacting factor where interactions were statistically significant.

## RESULTS

### Sample description

The analytic sample for research question 1 comprised 335,172 people alive with a recorded dementia diagnosis, and 3,065,858 people without dementia. For research question 2 and 3, the analytic sample comprised 2,083,297 individuals: 556,685 people with dementia and 1,526,612 matched controls (Figure 1).

**Figure 1.**
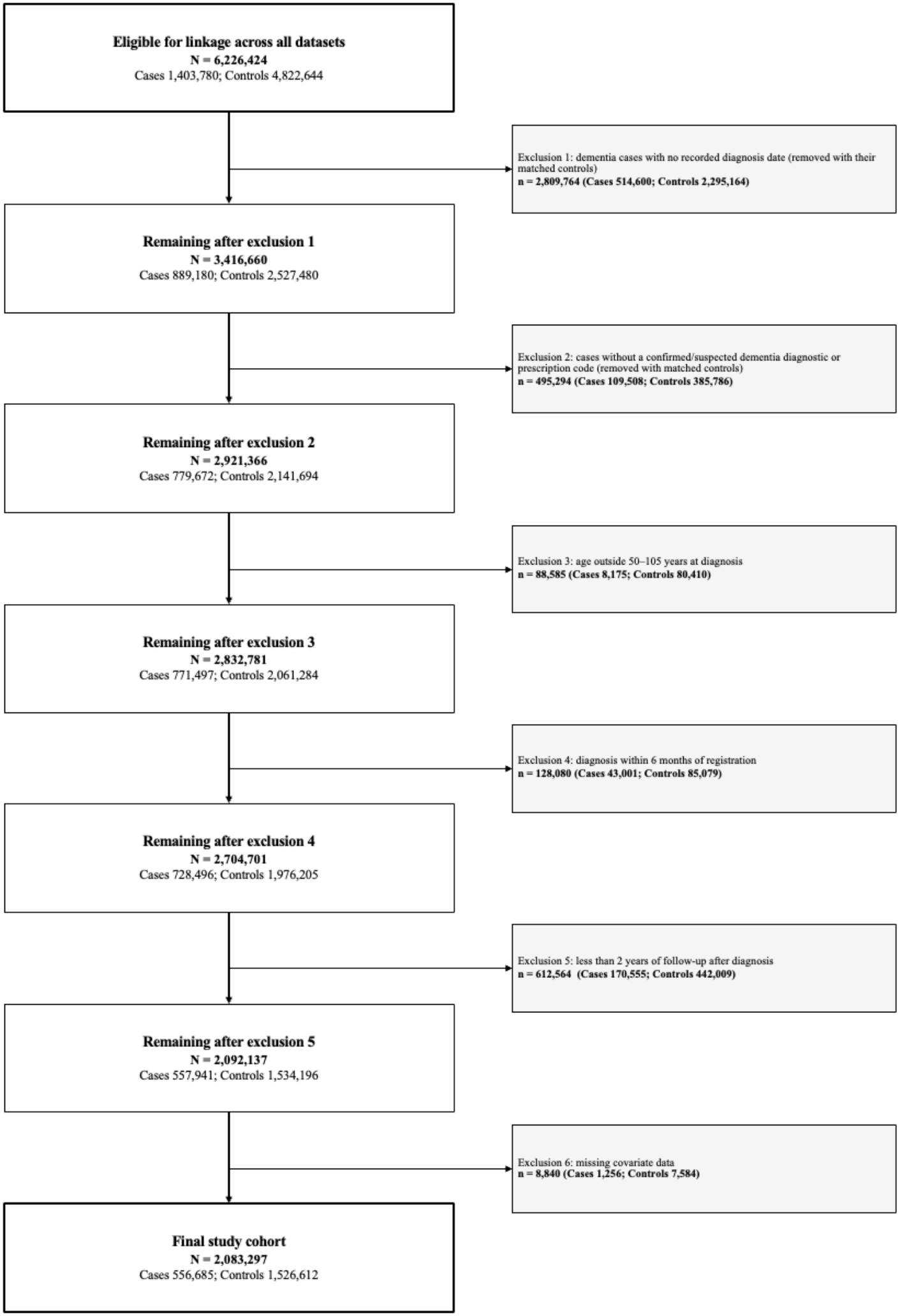
Study flow chart for research questions 2 and 3

Matching was performed before the exclusion of ineligible participants and a variable number of controls (up to 10) were retained per dementia cases; therefore the final analytical sample shows small residual differences in age and sex distribution, for which we controlled in adjusted models. For research questions 2 and 3, compared with controls, people with dementia were slightly older, more likely to be female, of White ethnicity, and have moderate or severe frailty, less likely to smoke, and had shorter follow-up (Table 1).

**Table 1.** Characteristics of the population at the time of entering the study (index date) for research questions 2 and 3.

|  |  | **Matched controls (1,526,612)** | **Dementia (556,685)** | **p-value** |
|---|---|---|---|---|
| **Age** |  | 73.90 (9.45) | 80.57 (8.75) | <0.001* |
| **Sex (Male)** |  | 694978 (45.5%) | 219533 (39.4%) | <0.001* |
| **Ethnicity** | Asian | 18456 (1.2%) | 3523 (0.6%) | <0.001* |
|  | Black | 28399 (1.9%) | 10506 (1.9%) |  |
|  | Mixed/Multiple | 6343 (0.4%) | 1684 (0.3%) |  |
|  | Other | 3751 (0.2%) | 540 (0.1%) |  |
|  | South Asian | 33384 (2.2%) | 8138 (1.5%) |  |
|  | Unknown | 72379 (4.7%) | 12946 (2.3%) |  |
|  | White | 1363900 (89.3%) | 519348 (93.3%) |  |
| **Smoking status** | Current smoker | 175326 (11.5%) | 44406 (8.0%) | <0.001* |
|  | Ex-smoker | 433861 (28.4%) | 156205 (28.1%) |  |
|  | Non-smoker | 797698 (52.3%) | 311504 (56.0%) |  |
|  | Missing data | 119727 (7.8%) | 44570 (8.0%) |  |
| **IMD** | Quintile 1 | 359498 (23.5%) | 119665 (21.5%) | <0.001* |
|  | Quintile 2 | 352788 (23.1%) | 123322 (22.2%) |  |
|  | Quintile 3 | 312791 (20.5%) | 114574 (20.6%) |  |
|  | Quintile 4 | 273203 (17.9%) | 104021 (18.7%) |  |
|  | Quintile 5 | 228332 (15.0%) | 95103 (17.1%) |  |
| **Rural-urban classification** | Urban | 1292355 (84.7%) | 475473 (85.4%) | <0.001* |
|  | Rural | 234257 (15.3%) | 81212 (14.6%) |  |
| **Follow-up duration** |  | 7.66 (4.82) | 6.49 (4.28) | <0.001* |
| **Severity of frailty** | No frailty | 1110482 (72.7%) | 263210 (47.3%) | <0.001* |
|  | Mild | 328455 (21.5%) | 190872 (34.3%) |  |
|  | Moderate | 74830 (4.9%) | 80312 (14.4%) |  |
|  | Severe | 12845 (0.8%) | 22291 (4.0%) |  |

*\*p<0.05*

Missing data on smoking status were associated with age, sex, ethnicity, median follow-up time, IMD, dementia status, and frailty status (all p < 0.001), suggesting data were not missing completely at random, and were therefore suitable for imputation.

### Research question 1: Frailty levels in people with and without dementia

Among people with dementia, 194,773 (58.1%) were living with frailty: 102,334 (30.5%) with mild frailty, 64,240 (19.2%) with moderate frailty, and 28,199 (8.4%) with severe frailty. Frailty was less common among people without dementia (χ^2^ =180,507, df=3, p < 0.001), of whom 902,040 (29.4%) were living with frailty: 664,075 (21.7%) mild, 192,259 (6.3%) moderate, and 45,706 (1.5%) severe. In both groups, frailty was more prevalent in women than men (dementia 61.7% vs 53.3%; controls 32.9% vs 26.0%) and increased markedly with age (Table 2).

**Table 2.** Frailty prevalence in England in 2021, by age and sex.

|  |  | **N** | **No frailty, n (%)** | **Mild, n (%)** | **Moderate, n (%)** | **Severe, n (%)** | **Any frailty, n (%)** |
|---|---|---|---|---|---|---|---|
| **Overall** | Dementia | 335,172 | 140,399 (41.9%) | 102,334 (30.5%) | 64,240 (19.2%) | 28,199 (8.4%) | 194,773 (58.1%) |
|  | No dementia | 3,065,858 | 2,163,818 (70.6%) | 664,075 (21.7%) | 192,259 (6.3%) | 45,706 (1.5%) | 902,040 (29.4%) |
| **By sex** |  |  |  |  |  |  |  |
| Male | Dementia | 144,006 | 67,192 (46.7%) | 41,870 (29.1%) | 24,799 (17.2%) | 10,145 (7.0%) | 76,814 (53.3%) |
|  | No dementia | 1,534,847 | 1,135,973 (74.0%) | 302,503 (19.7%) | 79,577 (5.2%) | 16,794 (1.1%) | 398,874 (26.0%) |
| Female | Dementia | 191,166 | 73,207 (38.3%) | 60,464 (31.6%) | 39,441 (20.6%) | 18,054 (9.4%) | 117,959 (61.7%) |
|  | No dementia | 1,531,011 | 1,027,845 (67.1%) | 361,572 (23.6%) | 112,682 (7.4%) | 28,912 (1.9%) | 503,166 (32.9%) |
| **By age group** |  |  |  |  |  |  |  |
| <60 | Dementia | 16,839 | 13,335 (79.2%) | 2,689 (16.0%) | 687 (4.1%) | 128 (0.8%) | 3,504 (20.8%) |
|  | No dementia | 294,079 | 278,073 (94.6%) | 14,659 (5.0%) | 1,254 (0.4%) | 93 (0.0%) | 16,006 (5.4%) |
| 60–64 | Dementia | 13,242 | 9,154 (69.1%) | 2,930 (22.1%) | 948 (7.2%) | 210 (1.6%) | 4,088 (30.9%) |
|  | No dementia | 232,902 | 207,705 (89.2%) | 22,315 (9.6%) | 2,628 (1.1%) | 254 (0.1%) | 25,197 (10.8%) |
| 65–69 | Dementia | 17,551 | 10,602 (60.4%) | 4,623 (26.3%) | 1,813 (10.3%) | 513 (2.9%) | 6,949 (39.6%) |
|  | No dementia | 314,113 | 263,103 (83.8%) | 43,693 (13.9%) | 6,452 (2.1%) | 865 (0.3%) | 51,010 (16.2%) |
| 70–74 | Dementia | 30,804 | 16,329 (53.0%) | 9,136 (29.7%) | 3,936 (12.8%) | 1,403 (4.6%) | 14,475 (47.0%) |
|  | No dementia | 514,035 | 396,677 (77.2%) | 97,619 (19.0%) | 17,189 (3.3%) | 2,550 (0.5%) | 117,358 (22.8%) |
| 75–79 | Dementia | 49,739 | 22,387 (45.0%) | 16,115 (32.4%) | 8,259 (16.6%) | 2,978 (6.0%) | 27,352 (55.0%) |
|  | No dementia | 658,914 | 455,640 (69.2%) | 160,577 (24.4%) | 36,386 (5.5%) | 6,311 (1.0%) | 203,274 (30.8%) |
| ≥80 | Dementia | 206,997 | 68,592 (33.1%) | 66,841 (32.3%) | 48,597 (23.5%) | 22,967 (11.1%) | 138,405 (66.9%) |
|  | No dementia | 1,051,815 | 562,620 (53.5%) | 325,212 (30.9%) | 128,350 (12.2%) | 35,633 (3.4%) | 489,195 (46.5%) |

*Any frailty = mild + moderate + severe frailty.*

### Research question 2: Impact of frailty on outcomes

#### Hospitalisations

Over the follow-up period (4.1 years in people with dementia and 6.8 years in matched controls), the median total bed days was 20 (IQR 2–56) for people with dementia, and 6 (IQR 0–26) in matched controls, in analyses that did not account for the shorter follow up time in the dementia group (Supplementary Table S4).

In analytic models that accounted for follow-up time, hospitalisation episodes increased with frailty severity in both groups. Total bed days showed a similar overall pattern, though among people with dementia this was driven only by the severe frailty category, with those with mild or moderate frailty having fewer bed days than those with no frailty (Table 3). The associations were significantly stronger in controls than people with dementia (interaction p<0.001); sensitivity analyses provided similar results (Supplementary Table S5).

**Table 3.** Unadjusted and adjusted associations between frailty and hospitalisation and mortality outcomes.

|  | **People with dementia** |  | **Matched controls** |  |
|---|---|---|---|---|
|  | **Unadjusted estimate (95% CI), p-value** | **Adjusted estimate (95% CI), p-value** | **Unadjusted estimate (95% CI), p-value** | **Adjusted estimate (95% CI), p-value** |
| **Hospitalisation episodes (IRR)** |  |  |  |  |
| Mild | 1.11 (1.10, 1.12), p<0.001* | 1.16 (1.15, 1.17), p<0.001* | 1.77 (1.76, 1.78), p<0.001* | 1.56 (1.55, 1.57), p<0.001* |
| Moderate | 1.26 (1.25, 1.27), p<0.001* | 1.33 (1.32, 1.35), p<0.001* | 2.63 (2.60, 2.66), p<0.001* | 2.18 (2.16, 2.21), p<0.001* |
| Severe | 1.37 (1.34, 1.39), p<0.001* | 1.47 (1.45, 1.50), p<0.001* | 3.74 (3.65, 3.84), p<0.001* | 3.07 (2.99, 3.14), p<0.001* |
| **Total bed days (IRR)** |  |  |  |  |
| Mild | 0.91 (0.90, 0.92), p<0.001* | 0.96 (0.95, 0.97), p<0.001* | 1.98 (1.96, 1.99), p<0.001* | 1.55 (1.54, 1.56), p<0.001* |
| Moderate | 0.93 (0.91, 0.94), p<0.001* | 0.99 (0.97, 0.99), p = 0.02* | 3.41 (3.36, 3.46), p<0.001* | 2.35 (2.32, 2.38), p<0.001* |
| Severe | 0.97 (0.95, 0.99), p = 0.03* | 1.04 (1.01, 1.07), p<0.001* | 5.10 (4.99, 5.27), p<0.001* | 3.28 (3.17, 3.38), p<0.001* |
| **All-cause mortality (HR)** |  |  |  |  |
| Mild | 1.24 (1.23, 1.26), p<0.001* | 1.09 (1.08, 1.10), p<0.001* | 2.19 (2.17, 2.22), p<0.001* | 1.42 (1.41, 1.43), p<0.001* |
| Moderate | 1.58 (1.56, 1.61), p<0.001* | 1.29 (1.27, 1.31), p<0.001* | 3.93 (3.87, 3.99), p<0.001* | 1.99 (1.96, 2.02), p<0.001* |
| Severe | 2.00 (1.96, 2.05), p<0.001* | 1.58 (1.54, 1.62), p<0.001* | 6.18 (6.01, 6.34), p<0.001* | 2.74 (2.66, 2.82) p<0.001* |

Reference category: no frailty. *\*p<0.05.*
Adjusted models control for ethnicity, IMD, rurality, smoking status, age, and sex, with random effects for practice.
IRR, incidence rate ratio; HR, hazard ratio

#### All-cause mortality

Of the 556,685 people with dementia, 465,055 (83.5%) died over follow-up (median 4.1 years), compared with 558,481 (36.6%) of 1,526,612 controls (median 6.8 years). In both groups, the median follow-up time was shorter with increasing frailty severity (Supplementary Table S6).

Schoenfeld tests indicated the proportional hazards assumption was violated, but because such tests detect even trivial deviations at large sample sizes^19^, and as we focused on the overall effect of frailty across follow-up, we used standard Cox regression, interpreting results as average hazard ratios over the follow-up period.

In both dementia and control groups, higher frailty severity was associated with increased mortality risk (Table 3). In line with our findings for hospitalisations, the association between frailty severity and mortality was stronger in controls than people with dementia (interaction p < 0.001). In adjusted models restricted to those with dementia, compared to people with no frailty, mortality risk increased with mild (HR 1.09, 95% CI 1.08–1.10, p < 0.001), moderate (HR 1.29, 95% CI 1.27–1.31, p < 0.001),and severe frailty (HR 1.58, 95% CI 1.54–1.62, p < 0.001). In the equivalent model for control participants, compared to those with no frailty, mild (HR 1.42, 95% CI 1.41–1.43, p < 0.001), moderate (HR 1.99, 95% CI 1.96–2.02, p < 0.001), and severe frailty (HR 2.74, 95% CI 2.66–2.82, p< 0.001) were associated with increased mortality risk. Results were unchanged in sensitivity analyses (Supplementary Table S5).

### Research question 3: Potential health inequalities

Table 4 summarises the interaction analyses, and stratified estimates are provided in Supplementary Materials (Supplementary Table S7 to S14). Among people with dementia, frailty more strongly predicted death in White individuals, those in the most deprived groups, and in rural areas, whereas in controls it more strongly predicted death in non-White individuals and, likewise, those in rural areas. Frailty was more strongly associated with hospitalisations in non-White than White individuals, in both groups.

**Table 4.** Results of interaction analyse.

|  | **White vs. Non-White** | **Most deprived vs. Least deprived** | **Urban vs. Rural** |
|---|---|---|---|
| **Hospitalisation episodes** | Dementia: p < 0.001*<br>Control: p < 0.001* | Dementia: p < 0.001*<br>Control: p < 0.001* | Dementia: p = 0.08<br>Control: p < 0.001* |
| **Total bed days** | Dementia: p < 0.001*<br>Control: p < 0.001* | Dementia: p = 0.51<br>Control: p < 0.001* | Dementia: p = 0.86<br>Control: p = 0.14 |
| **Mortality** | Dementia: p < 0.001*<br>Control: p < 0.001* | Dementia: p < 0.001*<br>Control: p < 0.001* | Dementia: p = 0.004*<br>Control: p = 0.001* |

*\*p<0.05*
*Direction of each significant interaction is reported in Supplementary Tables*

## DISCUSSION

We found that in England, almost 6 in 10 people with dementia were living with frailty, twice the proportion among control cases without dementia. A recent review similarly found a frailty prevalence of 66% using similar, deficit-accumulation indices^9^. In people without dementia, greater frailty was associated with more total bed days. Among people with dementia, those with mild or moderate frailty spent slightly fewer days in hospital than those without frailty, whereas those with severe frailty spent more days in hospital. The complexity of planning community discharge of people with dementia returning to their own homes, rather than a care home and with less care already in place could explain this. The Casey Commission has a remit to address “delayed discharges when care services don’t have capacity for those needing ongoing care”^20^.

Aligning with existing eFI studies in older adults^17^ and people with dementia^6^, we found a dose-response relationship between frailty and mortality, though this relationship was less strong in people with dementia. This may be a ceiling effect, as dementia already confers high baseline risk, or because more frequent healthcare contact allows earlier management of frailty-related deterioration in people with dementia.

Our findings on ethnicity and mortality were consistent with existing research showing that non-White older adults with dementia have a lower mortality risk than White individuals^10,21^. This may reflect survivor selection or a healthy-migrant effect – whereby people healthy enough to migrate, and survive to older age, are a resilient group^22^. Alternatively, frailty may be recorded less completely in minority ethnic groups, who tend to have less healthcare contact^23^. Our finding that frailty was more strongly associated with hospitalisations in non-White than White individuals, contrasts with previous studies in England, which have reported higher admissions among White, more deprived, and rural groups^11^. Overall, inequalities were more evident in controls, suggesting the more frequent healthcare contact of people with dementia may attenuate them.

People with dementia experienced hospitalisation and mortality outcomes at least as poor as those of the most severely frail people without dementia. This supports the approach of considering the needs of people with dementia and frailty together, as in the government’s planned Modern Service Framework^13^. It is notable that the 4 in 10 people with dementia who were not frail, had hospitalisation outcomes similar to the most severely frail people without dementia. People with dementia wish to be supported to live in the community for as long as possible^24,25^, and avoid unnecessary admissions^26^. However, they spent substantially more time in hospital than those without dementia at comparable frailty. Some of this time may be avoidable: those with little or no frailty but long admissions might be better supported through Hospital at Home or “virtual ward” models, though these models are not currently well set up for people with dementia (Wang et al). Around one in five people with dementia are first diagnosed during a hospital admission^27^ and earlier diagnosis could open routes to hospital avoidance and more timely community support.

Future research should explore what drives the relatively high hospital use among people with dementia who are not identified as frail, and which forms of community-based or preventative support are most effective for this group. As care shifts from hospital into the home, including through earlier discharge and Hospital at Home or virtual ward models, the role and capacity of unpaid carers also warrant investigation: the feasibility and impact of these approaches are likely to depend considerably on the formal and informal support available.

### Limitations

We used the original eFI as it was the most extensively validated frailty index in UK primary care. However, it does not capture conditions severities, nor whether a recorded condition has since been treated or resolved, and it weights all deficits equally; a revised version (eFI2)^28^ that applies deficit weighting would have been an alternative approach. Dementia may contribute to the eFI both directly (via a dementia diagnosis) and indirectly (via the eFI’s “memory and cognitive problems” deficit), so people with dementia will by definition have a slightly higher eFI than they would without the diagnosis. Electronic health record data quality varies with GP recording practices and missing data may affect generalisability. Some control participants will have had undiagnosed dementia; this may have attenuated the observed differences. Finally, because ascertainment relies on healthcare contact, frailty may be recorded more completely in people with dementia, while individuals less engaged with services – including some minority ethnic and more deprived groups – may be underrepresented, limiting the precision of subgroup estimates.

## Conclusion

Over half of people with dementia in England were living with frailty. Greater frailty severity was associated with higher rates of hospitalisation and mortality, with similar but weaker associations than in controls, and its impact varied by health inequalities. These findings highlight the need to identify and manage frailty as a routine part of dementia care, attentive to the differing needs of population subgroups. People with dementia without frailty are a group at high risk of long hospital admissions; interventions to avoid or shorten admission could have an important impact on the ability of this group to remain living in the community for as long as possible, so their inclusivity to people with dementia should be prioritised.

## Data Availability

The data used in this study were obtained from the Clinical Practice Research Datalink (CPRD) under licence and cannot be shared by the authors. Data are available from CPRD (https://www.cprd.com) subject to protocol approval and the relevant licence agreements.

## Funding statement

This research is funded through the NIHR Policy Research Unit in Dementia and Neurodegeneration – Queen Mary University of London, reference NIHR206110. The views expressed are those of the author(s) and not necessarily those of the NIHR or the Department of Health and Social Care.

## Notes

### Competing Interest Statement

The authors have declared no competing interest.

